# Estimating the sleep period time window based on a hip-worn accelerometer collected in children and adults

**DOI:** 10.1101/2025.11.25.25340956

**Authors:** Jairo H Migueles, Vincent T van Hees, Michael J Stein, Michael F Leitzmann, Hansjoerg Baurecht, Claas Lendt

## Abstract

**Background:** Accurately detecting the Sleep Period Time (SPT) window in the daily life is essential for understanding habitual sleep and health. Although actigraphy devices (accelerometers) placement varies across studies, most SPT-detection algorithms are developed for wrist data. Open-source algorithms support reproducibility and transparency in estimating the SPT.

**Aims:** To optimise and evaluate two open-source algorithms, HDCZA and HorAngle, for estimating the SPT window using hip-worn accelerometer data.

**Methods:** A total of 109 children and 194 adults wore wrist and hip accelerometers for six nights and completed sleep diaries. An established algorithm combining wrist and diary data served as the reference. HDCZA and HorAngle parameters were optimised using Bayesian optimisation on 60% of the sample and evaluated in the remaining 40%.

**Results:** Mean differences for sleep onset and wake-up were –3 and 4 minutes for HDCZA (limits of agreement [LoA]: −221,215 and −185,194; root-mean square error [RMSE]=111 and 97) and 0 and −4 minutes for HorAngle (LoA: −199,199 and −223,214; RMSE=111 and 112). For SPT duration, mean differences were 7 minutes (LoA: −252,266; RMSE=132) for HDCZA and −4 minutes (LoA: −254,246; RMSE=128). No significant differences in SPT duration were found (P=0.774; P=0.237). Both algorithms showed moderate agreement with the reference in ranking sleep duration (κ ≈ 0.56−0.58). Differences were unrelated to age or sex but linked to non-wear time.

**Conclusions:** Both open-source algorithms demonstrated value for estimating the SPT window from hip data. While HDCZA requires no additional sensor-specific parameters, HorAngle depends on accurate axis identification.

**Statement of Significance:** Accurately estimating the sleep period time (SPT) window from hip-worn accelerometers is essential for studies assessing sleep in free-living conditions. However, most available algorithms were developed for wrist-worn data. This study optimised and validated two open-source algorithms, HDCZA and HorAngle, for hip-worn accelerometer data in children and adults. Both algorithms performed comparably to a wrist-based reference using sleep diaries, showing consistent agreement across age and sex. These methods enable researchers to estimate habitual sleep without additional sensors or diaries, improving reproducibility and scalability in observational research. The algorithms are openly implemented in the GGIR R package, offering accessible and standardised tools for analysing hip-based accelerometer data.

## Introduction

The sleep period time (SPT) window refers to the time between sleep onset and the end of sleep.^1^ This period is a key metric for studying sleep in both clinical and population-based studies. Major health organisations, including the American Academy of Sleep Medicine and the World Health Organisation, base their sleep duration recommendations on the SPT window.^2–4^

Wrist-worn actigraphy is widely used for estimating the SPT window in the daily life of study participants.^5^ However, a number of population-based cohorts have adopted hip-worn accelerometers for 24-hour activity monitoring. Examples include as NAKO (Germany),^6^ Tromsø (Norway),^7^ ELSA (Brazil),^8^ Raine (Australia),^9^ and OPACH (United States),^10^ with a combined sample size of >85,000 participants with valid accelerometer data across a wide age range. All these studies store unprocessed ‘raw’ acceleration values in gravitational units, as opposed to pre-processed ‘count’ values. Further, most of these studies did not employ a sleep diary, which necessitates relying solely on actigraphy to detect the SPT window. The advantages of raw value collection are increased analytical freedom and the ability to re-use algorithms across sensor brands. This form of actigraphy is commonly referred to as accelerometry in reference to the acceleration sensor.

No algorithms have been reported in the literature for classifying the SPT window from raw hip-worn accelerometer data. However, open-source R package GGIR as widely used for wrist-worn accelerometer data offers two algorithms that have not been optimized and evaluated yet for data from the hip.

In order to optimize and evaluate algorithms for accelerometer data a reference method is needed that can consider the full 24 hour window.^11^ Although polysomnography (PSG) is the gold standard for classifying sleep, it is not feasible to be worn continuously over a measurement period of multiple hours or days. As a result, whether short naps near nighttime, such as an evening nap in front of the television, are included in the main sleep period depends on when participants choose to wear the PSG, rather than on a standardized methodology. Instead, a method based on wrist-worn accelerometer data, guided by a sleep diary, offers a reference method that captures 24-hour sleep-wake patterns over multiple days, something not feasible with PSG.

In this study, we aimed to optimise and evaluate two existing algorithms^12^ for hip-worn accelerometer data in children and adults by using a reference method that detects SPT window based on wrist-accelerometer data guided by sleep diary.

## Methods

### Study Population

The data for this study were derived from the baseline assessments of two clinical trials: ActiveBrains^13^ and ACTIBATE trials.^14^ The ActiveBrains trial included 109 children (41% girls) aged 8−12 years with overweight or obesity, while the ACTIBATE trial enrolled 194 sedentary, healthy, young adults (65% women) aged 18−35 years. The data were split into an optimization and a test set by taking a random subset of 60% and 40% from each cohort (n = 181 and n = 122, respectively).

Written informed consent was obtained from all participants in the ACTIBATE trial and from the parents or legal guardians of the children in ActiveBrains. Both trials were approved by the Ethics Committee of the University of Granada and were registered at ClinicalTrials.gov (ActiveBrains: NCT02295072; ACTIBATE: NCT02365129).

### Data collection and pre-processing

In both studies, participants were instructed to wear two ActiGraph GT3X+ accelerometers (ActiGraph, Pensacola, Florida, US) simultaneously for seven consecutive days and six nights, one on their right hip and one on their non-dominant wrist. Throughout the monitoring period, participants were instructed to maintain a sleep diary, recording their sleep onset and wake-up times. All accelerometers were configured to collect data at a 100 Hz sampling frequency, with the ActiGraph’s idle sleep mode disabled.

The raw acceleration data were downloaded via the ActiLife v6.13.3 software (ActiGraph, Pensacola, Florida) into .gt3x files and processed using the open-source R package GGIR v3.2-7.^12^ We applied the default GGIR data pre-processing steps, including: (i) auto-calibration of the raw accelerations; (ii) detection of non-wear time; (iii) estimation of the angles of each accelerometer’s axis relative to the horizontal plane per 5-second epoch.

After processing, the GGIR sleep output data were filtered to retain only nights for which sleep diaries were available. Additionally, nights for which more than one-third of the sleep window was marked as invalid (e.g., non-wear time) in either the wrist or hip data were excluded from all analyses.

### Sleep Period Time window estimation

The estimation of the SPT window in GGIR is estimated in three stages:

1. Detection of rest periods, which in GGIR are referred to as Sustained Inactivity Bouts (SIB). For this, we used the algorithm proposed by van Hees et al. (2015),^15^ which identifies periods of ‘timethreshold’ minutes in which the angle of the z-axis does not change by more than ‘anglethreshold’ degrees.
2. Identification of time windows that guide the eventual sleep detection, referred to as guider window. The methods used to define the guider window are referred to as ‘guiders’. The following guiders are used in this study:

- **Sleep log:** Sleep onset and wake-up time as reported by study participants in their sleep diary.
- **HDCZA:** This algorithm was designed for studies with wrist-worn accelerometer data in which no sleep log is available.^1^ The algorithm consists of nine steps as discussed elsewhere.^1^ In short, steps 1-6 attempt to classify time periods with limited change in the angle of the accelerometer’s z-axis. In the present study we have simplified step 6 with a single threshold represented by GGIR parameter ‘HDCZA_threshold’. Next, step 7 extracts rest blocks longer than the number of minues specified with parameter ‘spt_min_block’. Step 8 merges rest blocks separated by gaps shorter than the number of minutes specified with parameter ‘spt_max_gap_dur’ and with gap/rest ratio lower than the ratio specified with parameter ‘spt_max_gap_ratio’. Step 9 looks for the longest resulting rest block in the day, which represents the guider window.
- **HorAngle:** This represents a new algorithm designed for hip-worn accelerometer data. The algorithm searches for the longest period during which the longitudinal axis of the accelerometer is horizontal, which is used as a proxy for a horizontal orientation of the trunk. For this, we specified parameter ‘longitudinal_axis = 2’ to indicate that for the ActiGraph brand the second axis represents the longitudinal axis. The algorithm then identifies when this axis is horizontal, i.e., when it is between -*X* and +*X* degrees, where *X* is specified with parameter ‘HorAngle_threshold’. Finally, the algorithm uses the classified periods of being horizontal as input for the same final 3 steps (steps 7 to 9) as the HDCZA algorithm to find the main window to act as guider.
3. Identification of the overlap between the windows identified in steps 1 and 2 to define the SPT window. Briefly, SIBs overlapping with the guider window are considered to be part of the SPT, with the first and last overlapping SIBs indicating the sleep onset and wake-up times.

As our aim is to optimise the estimation of the SPT-window rather than the SIBs, we configured GGIR to treat the guider window as the final estimate for SPT (i.e., with GGIR parameters anglethreshold = 100, timethreshold = 1, ignorenonwear = FALSE, relyonguider = TRUE).

All parameter definitions can be found in Table 1.

**Table 1:** Definition of parameters, configuration in the referent method (wrist accelerometers guided by sleep logs) and parameters’ bounds used for the optimisation of the SPT estimation algorithms.

| Parameter | Description | Wrist and sleep log (ref) | Hip and HDCZA | Hip and HorAngle |
| --- | --- | --- | --- | --- |
| anglethreshold | Angle threshold (degrees) for SIBs detection. | 5 | 100 | 100 |
| timethreshold | Time threshold (minutes) for SIBs detection. | 5 | 1 | 1 |
| HDCZA_threshold | Threshold used by the HDCZA algorithm to define candidate blocks for the guider window. | - | 0.001 to 0.05 or 0.13 to 0.7 | - |
| HorAngle_threshold | Threshold used by the HorAngle to define when the trunk is in horizontal position. | - | - | 40 to 70 |
| spt_min_block | Minimum size of the block (minutes) to be considered a candidate for the guider window. | - | 10 to 60 | 10 to 60 |
| spt_max_gap_dur | Maximum gap allowed (minutes) between candidate blocks to be merged as a combined into a single block. | - | 20 to 90 | 20 to 90 |
| spt_max_gap_ratio | Maximum ratio between gap duration and either rest block next to it. | - | 0 to 1 | 0 to 1 |

SPT: sleep period time; SIB: sustained inactivity bout; HDCZA: Heuristic algorithm looking at Distribution of Change in Z-Angle.

### Parameter optimisation

Wrist accelerometer data guided by sleep diaries were used as reference method as this approach has previously been demonstrated to provide acceptable validity^1^ and combines the strengths of both sleep log and wrist accelerometry.^15^.

The performance metric used during optimisation was the average root mean square error (RMSE) between the estimated sleep onset and wake-up times provided by the algorithm and the reference values.

#### Optimisation procedure

The full data set was split into a training set (n = 181, 60%) and a test set (n = 122, 40%), ensuring proportional representation from the ActiveBrains and the ACTIBATE studies.

First, we used the training set to compare two combinations of search ranges for the ‘HDCZA_threshold’ parameter as used by the HDCZA algorithm in a 3-fold cross-validation in the full training set (fold size was 1/3 of the training set), see Table 1. This was done to narrow the search space and to reduce the risk of overfitting. To identify the most robust set of parameter ranges we used the product of the mean RMSE and standard deviation of the RMSE across folds as a selection criterion to balance both average performance and stability across data splits. Next, the most robust set of search ranges were used for the final optimisation.

The parameters for both HDCZA and HorAngle were then optimised in the full training set and the test set was used for the final evaluation of model performance. Here, the parameters were optimised using the “ParBayesianOptimization” R package for Bayesian optimisation.^16^ This was configured to use 8 initial points and 20 iterations for the cross-validation as used for HDCZA and 10 initial points and 50 iterations for training the final model as used for both algorithms. Further, the expected improvement acquisition function was selected to guide the optimisation.

The search space for each algorithm parameter can be found in Table 1.

## Statistics

Participant characteristics were summarised using means and standard deviations. We evaluated the performance of two algorithms, HDCZA and HorAngle, using their respective optimised parameters.

Algorithm performance was evaluated as follows. First, agreement of the algorithms with the reference values was assessed through Bland–Altman plots and the RMSE. To explore the shape of the distribution, we quantified the proportion of nights with absolute errors exceeding 0.5, 1, 2, and 5 hours. In addition, we investigated potential sources of error by fitting linear mixed-effects models, with estimation errors (for sleep onset and wake-up time) as dependent variables. The fixed effects included age group, sex, day type (weekday or weekend), and levels of non-wear time in hip or wrist accelerometer data. Participant ID was modeled as a random intercept to account for repeated measures. These models were fitted using the lmer function from the lmerTest R package.^17^ To evaluate whether the algorithms preserved the relative ranking of individuals by SPT window duration, we categorized reference and estimated SPT durations into quintiles and calculated squared-weighted Cohen’s kappa with corresponding confusion matrices.

To assess the added value of optimisation, we also evaluated the HDCZA algorithm using the default settings as implemented in GGIR, which were originally designed for wrist-worn accelerometer data.

## Results

The study included a total of 303 participants, comprising 109 children (mean age: 10.0 [SD=1.1] years; 44 girls) and 194 adults (mean age: 22.1 [SD=2.2] years; 124 women).

### Parameter optimisation and cross-validation

#### HDCZA algorithm

The results from the 3-fold nested cross-validation for the exploration of the appropriate search space for such algorithm are presented in Table S1 (Supplement). The second bound set resulted in a lower mean × standard deviation of RMSE (0.093 vs. 0.060). Therefore, it was selected for the final optimisation of the HDCZA algorithm using the full training set.

The Bayesian optimisation of the HDCZA algorithm resulted in the following optimal parameter values: HDCZA_threshold = 0.259, spt_min_block_dur = 34, spt_max_gap_dur = 52, and spt_max_gap_ratio = 0.282, see Figure 1.

**Figure 1:**
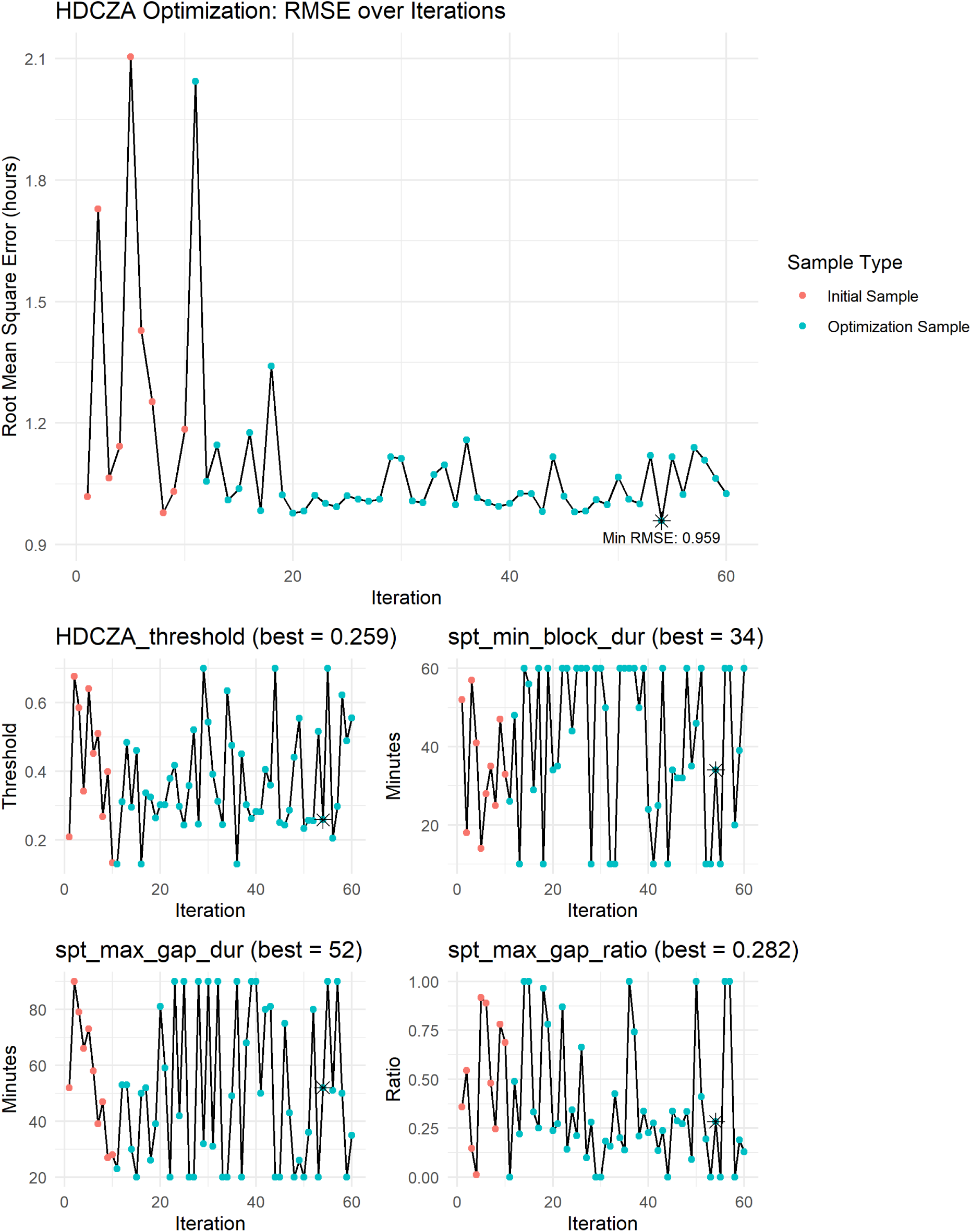
Optimisation output of parameters for the HDCZA algorithm. Star indicates the optimized value.

#### HorAngle algorithm

The Bayesian optimization of the HorAngle algorithm resulted in the following optimal parameter values: HorAngle_threshold = 58, spt_min_block_dur = 60, spt_max_gap_dur = 20, and spt_max_gap_ratio = 0.034, see Figure 2.

**Figure 2:**
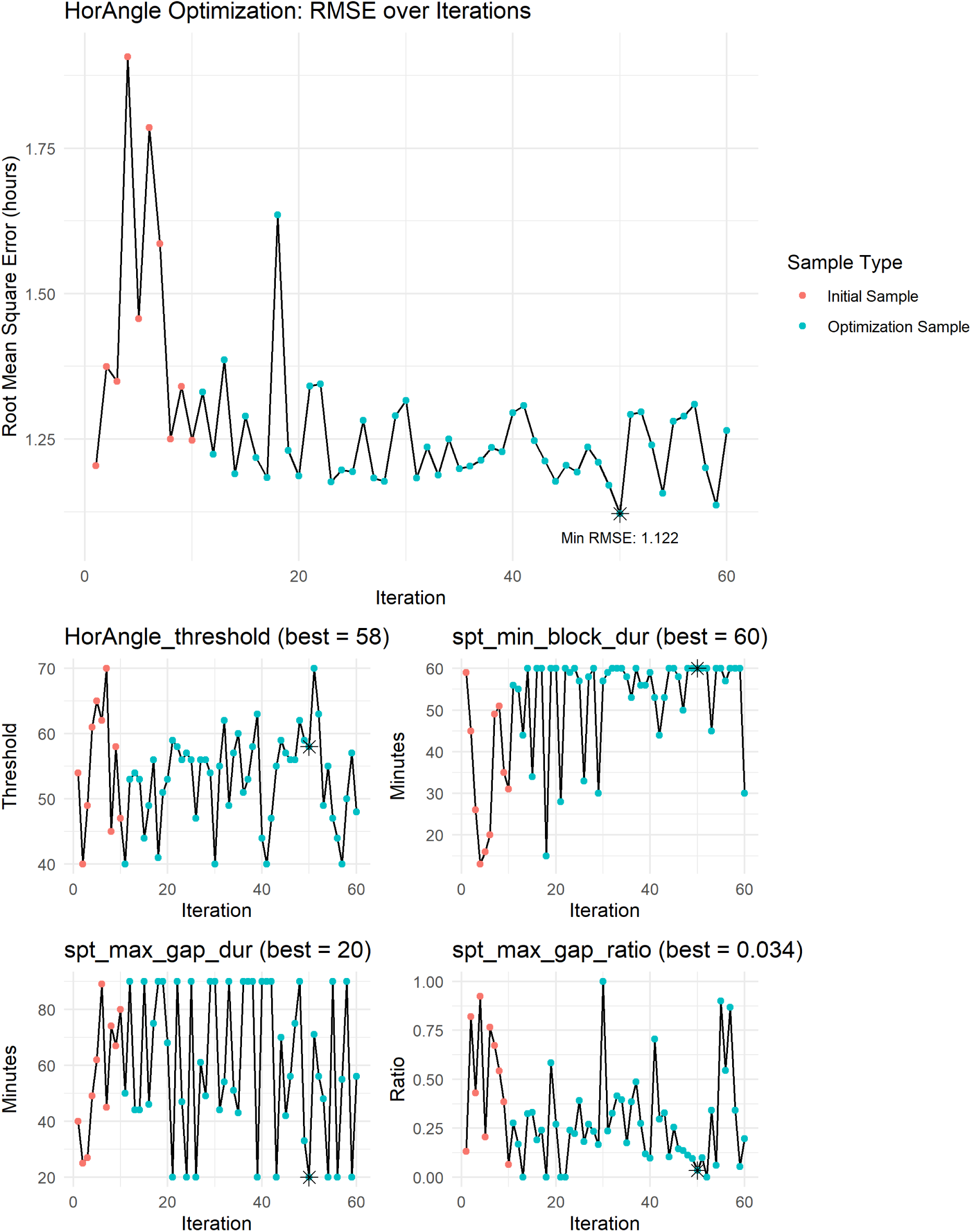
Optimisation output of parameters for the HorAngle algorithm. Star indicates the optimized value.

### Evaluation of the algorithms

For sleep onset, the HDCZA showed a mean difference of −3 minutes (*P* = 0.741) with limits of agreement (LoA) from −221 to 215 minutes, and with a RMSE of 111.1. The HorAngle algorithm performed similarly, with a mean difference of 0 minutes (LoA: −199 to 199 minutes, *P* = 0.967), and RMSE of 101.4, see top row of Figure 3.

**Figure 3:**
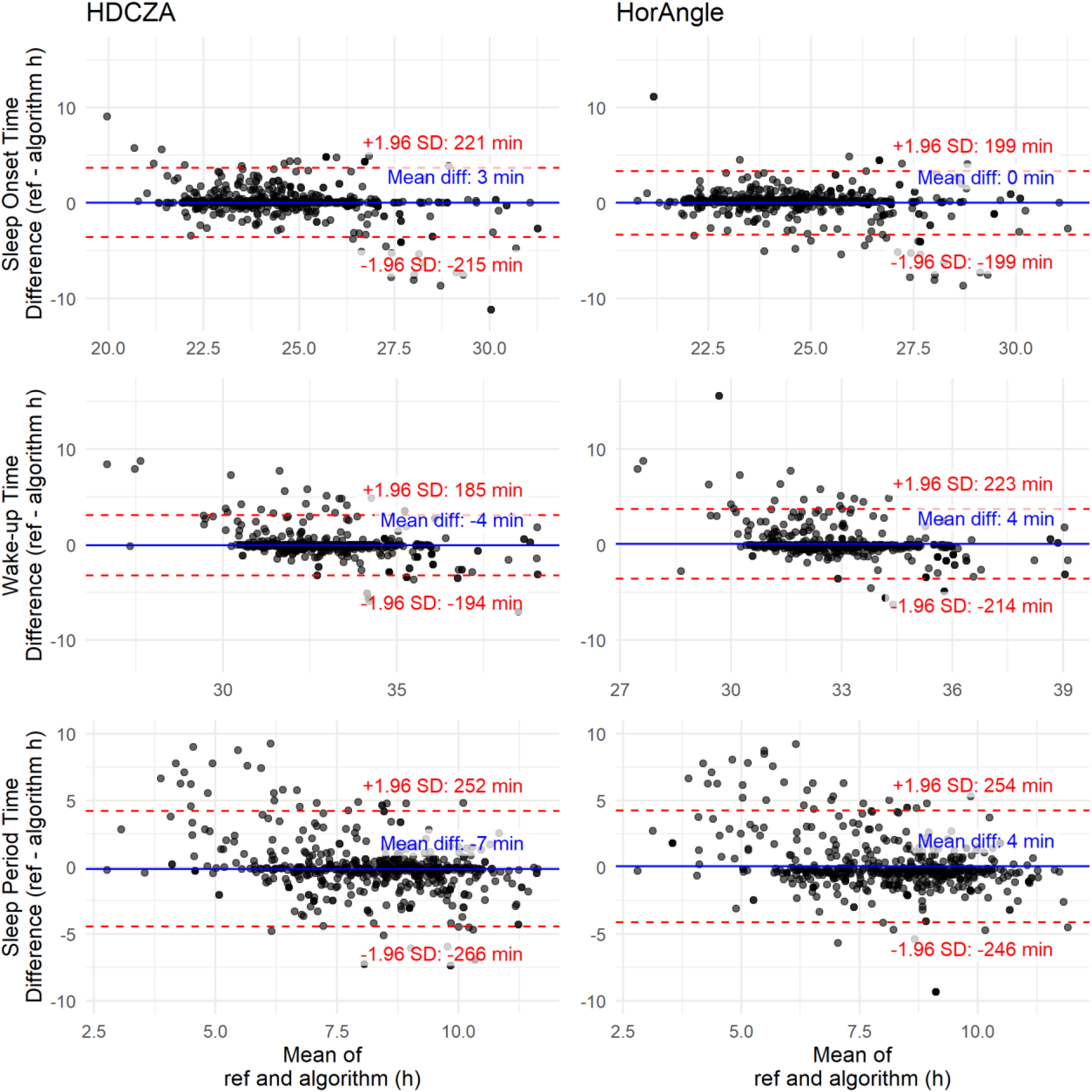
Bland Altman plots for the comparison of algorithm-estimated sleep onset time, wake-up time, and SPT with reference values (ref).

For wake-up time, the optimised HDCZA showed a mean difference of 4 minutes (LoA: −185 to 194, *P* = 0.537), and a RMSE of 96.7. The HorAngle showed a similar performance (−4 minutes; LoA: −223 to 214; *P* = 0.536; RMSE = 111.5), see middle row of Figure 3.

In terms of SPT window duration, the HDCZA yielded a mean difference of 7 minutes (LoA: −252 to 266; *P* = 0.324; RMSE = 132.4), while the HorAngle algorithm showed comparable results (-− minutes; LoA: −254 to 246; *P* = 0.557; RMSE = 127.6), see bottom row of Figure 3.

The HDCZA and HorAngle algorithms both resulted in a similar proportion of nights falling within the lowest error threshold (<60 minutes), 72.3% and 74.4% respectively, Table 2. Classification of participants into quintiles of SPT duration showed moderate agreement with the reference (weighted κ = 0.56 for HDCZA and 0.58 for HorAngle, p < 0.001), with most misclassifications occurring between adjacent quintiles, see Table S2 and Table S3 in the Supplement.

**Table 2:**
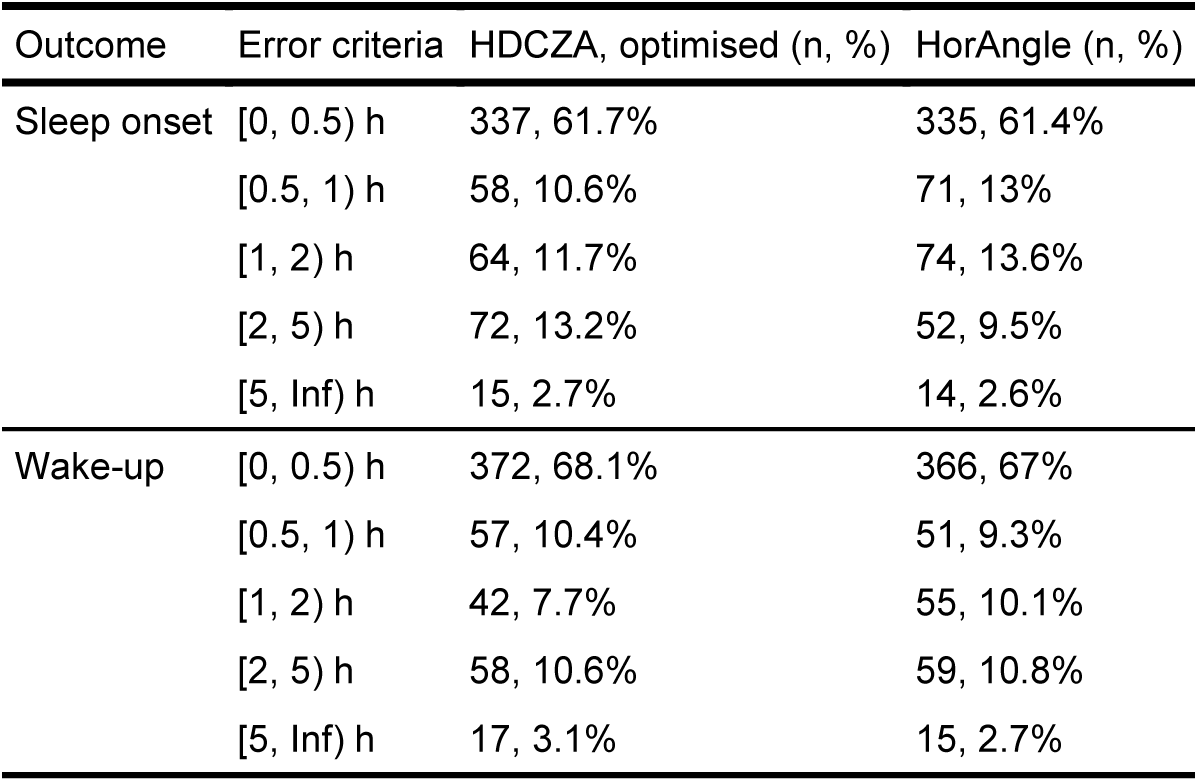
Distribution of nights classified by the error committed by each algorithm in the detection of sleep onset and wake-up times.

Examples of SPT window estimates from the HDCZA and the HorAngle algorithms can be found in the Supplementary Material (Figures S1 to S3).

#### Effect of potential confounders on the performance of the algorithms

For sleep onset, none of the covariates were significantly associated with differences between reference method and the HDCZA estimate, see Supplementary Table S4. However, in the HorAngle model, the presence of a non-wear mismatch (i.e., non-wear detected on only one device) was associated with a significantly earlier onset estimate (β = −0.740, P = 0.002), indicating potential sensitivity to inconsistent device wear time, see Supplementary Table S4.

For wake-up time, several variables were significantly associated with differences between the reference method and the HDCZA estimates, including: Child age group (β = −0.366, P = 0.016), female sex (β = −0.419, P = 0.003), hip non-wear (moderate) (β = −0.848, P = 0.011), and hip non-wear (high) (β = −1.698, P = <0.001). In contrast, for HorAngle only wrist non-wear showed a significant association (β = −1.337, P = 0.017), see Supplementary Table S4.

#### Secondary analysis: default HDCZA performance

In a secondary analysis, we evaluated the performance of the default HDCZA to explore the added value of optimisation (Figure S4). For sleep onset, the mean difference was −24 minutes (LoA: −273 to 224), for wake-up time 19 minutes (LoA: −203 to 241), and for SPT duration 43 minutes (LoA: −237 to 323).

## Discussion

This study proposes two candidate algorithms to estimate the SPT window from hip-worn accelerometers as widely used by several large cohort studies around the world. Firstly, we optimised the parameters for the HDCZA algorithm, originally designed for wrist-based data, and we introduced and optimised the HorAngle algorithm, which leverages posture-related information to estimate the SPT window. Both approaches were evaluated against a reference combining wrist-worn accelerometers and sleep diaries, which has demonstrated value in the previous studies.^15^ Our findings show similar performance of these two algorithms to estimate the SPT window based on hip accelerometer data, offering valuable alternatives in studies using 24-hour hip accelerometer data monitoring.

Although we refer to “sleep” throughout this study, it is important to note that we are not measuring sleep in the physiological or neurological sense. Our methods estimate the SPT window based on movement patterns, not direct indicators of sleep such as those captured via PSG. This distinction is critical, particularly in the context of free-living conditions, where neurological measurements are not feasible and no true gold standard for SPT detection exists.^11^

The HorAngle algorithm, which leverages the angle of the longitudinal axis as a proxy for trunk orientation during sleep, performed comparably or slightly better than the optimised HDCZA in some metrics, suggesting it is particularly better suited for hip data. However, a potential limitation is that HorAngle depends on the accurate identification of the accelerometer axis aligned with the body’s longitudinal axis. The intended axis to align with the longitudinal axis of the body is usually specified by the manufactured as the Y axis. However, this may vary across accelerometer brands and, in some studies the orientation of attachment is not properly controlled, for example when participant are instructed to carry the accelerometer in a pocket. Further, body shape and clothing may affect the orientation of the axis Y of the accelerometer relative to the longitudinal axis.

The proportion of nights showing large differences from our reference values (>2 hours) was comparable between the HDCZA and HorAngle algorithms. Linear mixed models indicated consistent performance across both children and adults, suggesting that the algorithms are applicable across age groups and sexes. However, these models also identified non-wear time as a source of residual error, which was associated with poorer accuracy in specific models. This highlights the need for further research into effective strategies for handling missing data due to non-wear, including the development of suitable imputation methods.

Previous efforts to estimate the SPT window using hip-worn accelerometers have relied primarily on algorithms developed for proprietary activity count data. Some examples include work by Tudor-Locke et al. (2014),^18^ Tracy et al. (2014),^19^ and McVeigh et al. (2016),^11^ which applied rule-based approaches to identify sleep intervals based on thresholds or changes in activity counts. Although the underlying algorithm to derive these activity counts has been published,^20^ we chose not to implement these methods in the present analysis for two reasons. First, fair method comparisons require testing in independent datasets rather than re-optimizing or re-applying algorithms on the same data used for their development. Second, we prioritized approaches based on raw accelerometer metrics (e.g., acceleration in gravitational units, posture angles), which are more interpretable and generalizable than abstract count units. Direct numerical comparison with prior findings is also limited by differences in reference standards. The earlier studies evaluated algorithms against visual inspection of accelerometer traces, which carries a risk of correlated errors because the same data source informs both algorithm and reference. By contrast, our study relied on an independent criterion measure, which provides a more robust assessment of validity.

More recent applications have further developed, tested, or adapted the algorithms introduced in the previous paragraph^11,18,19^ in diverse populations.^21–23^ When comparing our results to these previous studies, the performance of our optimized algorithms appears competitive. For instance, the HDCZA algorithm, when applied to hip-worn Actigraph data in children by Meredith-Jones et al., showed a mean difference of −6 (95% CI: −15, 4) minutes for sleep onset, −11 (95% CI: −40, 18) minutes for sleep offset, and 8 (95% CI: −5, 31) minutes for SPT-window duration.^24^ For Axivity devices worn on the back, Meredith-Jones et al.^24^ reported a mean difference of −21 (95% CI: −37, −5) minutes for sleep onset, −8 (95% CI: −32, 17) minutes for sleep offset, and 14 (95% CI: −8, 35) minutes for SPT-window duration using HDCZA. Likewise, the Count-Scaled (CS) algorithm, when used with Actigraph hip data, showed a mean difference of −12 (95% CI: −20, −4) minutes for sleep onset, 16 (95% CI: 1, 31) minutes for sleep offset, and 28 (95% CI: 13, 43) minutes for SPT duration. With Axivity devices on the back, the CS algorithm reported −20 (95% CI: −28, −13) minutes for sleep onset, 5 (95% CI: −10, 20) minutes for sleep offset, and 25 (95% CI: 10, 41) minutes for SPT duration. Our optimised HDCZA and HorAngle algorithms show better mean differences and CIs for sleep onset (HDCZA: −3 [95% CI: −12, 7]; HorAngle: −0 [95% CI: −9, 8]), wake-up (HDCZA: 4 [95% CI: −4, 13]; HorAngle: −4 [95% CI: −14, 5]), and SPT-window duration (HDCZA: 7 [95% CI: −4, 18]; HorAngle: −4 [95% CI: −15, 7]) compared to these. However, comparisons across studies are often limited due to differences in study design, reference methods, and outcome metrics.

Some other implementations have explored machine learning to classify sleep versus wake states, albeit not specifically focused on the SPT window.^25^ Although performance was improved significantly with random forests compared with conventional heuristic approaches, this gain often comes at the expense of interpretability. For example, a recent study by Weitz et al.^26^ applied a machine learning model to estimate time in bed using hip-worn accelerometers. While this illustrates growing interest in automating sleep-related estimates from hip accelerometer data, it shares similar limitations in terms of interpretability. Moreover, their use of chest-worn data for visual annotation may introduce correlated measurement error with hip-worn devices given that both are attached to the same part of the body, and the rationale for visually defining time in bed is not clearly justified. In contrast, our approach balances interpretability and performance by combining domain-informed parameter structures with data-driven Bayesian optimisation. This enables fine-tuning while retaining transparency and behavioral plausibility in the algorithm.

### Implications

Both the HorAngle and HDCZA algorithms are implemented in the permissively licensed open-source GGIR R package,^12^ ensuring compatibility with established GGIR pipelines already used in large epidemiological cohorts (e.g., NAKO, ELSA, Tromsø). This facilitates direct application of our methods within existing workflows and supports replication across diverse datasets. Additionally, the code and materials for the application of the Bayesian optimisation, and all the analyses conducted for the present study are available in an open-source GitHub repository (https://github.com/jhmigueles/spt_detection_hip).

Our findings have implications for sleep research in epidemiological cohorts using hip-worn accelerometers. As seen in large-scale cohort studies, hip placement is frequently chosen for 24-hour physical activity monitoring, yet existing sleep estimation methods for wrist-worn accelerometer data are suboptimal when applied to such data. This study provides validated, open-source methods that can improve the extraction of sleep metrics without requiring auxiliary information like sleep diaries, thus increasing scalability and reproducibility while decreasing participant burden. Example GGIR function calls using the validated optimised parameters are available in the Supplement.

### Strengths and Limitations

A strength of this study is the evaluation under free-living conditions which come with the challenge that rest period not part of the actual SPT window need to be separated. Furthermore, the use of two age groups (children and young adults) enhances generalisability. We also employed a rigorous optimisation approach involving nested cross-validation and Bayesian methods to prevent overfitting. Importantly, both algorithms evaluated in this study are implemented as open-source tools within the GGIR R package. Limitations include the relatively small and homogeneous sample compared to national cohorts, which may limit applicability to older adults or populations with sleep disorders. Noteworthy, these algorithms infer only the SPT window and do not classify sleep as such; for sleep detection, future research is needed using PSG as reference.

## Conclusion

We identified two candidate algorithms for estimating the SPT window. The novel HorAngle algorithm performed similarly to the HDCZA algorithm, providing a valuable alternative in settings where hip-based monitoring is used. Importantly, while the HDCZA algorithm can be applied directly without sensor-specific configuration, the HorAngle algorithm relies on the assumption that accelerometer is well aligned with the body’s longitudinal axis. Nevertheless, further evaluation of these parameters in independent samples is recommended to confirm generalisability and clinical relevance.

## Data Availability

All data produced in the present study are available upon reasonable request to the authors.

## Funding

The ActiveBrains project was supported with grants DEP2013-47540, DEP2016-79512-R, DEP2017-91544EXP, and RYC-2011-09011 from the Spanish Ministry of Economy and Competitiveness and the European Regional Development Fund and by grant PID2020-120249RB-I00 from the MCIN/AEI/10.13039/501100011033. Additional funding was obtained from the Andalusian Operational Programme supported with grant B-CTS-355UGR18 from the European Regional Development Fund. The ACTIBATE project was supported by the Spanish Ministry of Economy and Competitiveness via Fondo de Investigación Sanitaria del Instituto de Salud Carlos III (PI13/01393) and PTA 12264-I, Retos de la Sociedad (DEP2016-79512-R), and European Regional Development Funds (ERDF), by the Spanish Ministry of Education (FPU13/04365, FPU15/04059, FPU16/05159, FPU16/02828, FPU17/01523 and FPU19/01609), the Fundación Iberoamericana de Nutrición (FINUT), the University of Granada Plan Propio de Investigación 2016-Excellence actions: Unit of Excellence on Exercise and Health (UCEES), AstraZeneca HealthCare Foundationand by the Junta de Andalucía, Consejería de Conocimiento, Investigación y Universidades (ERDF, SOMM17/6107/UGR), the Junta de Andalucía, Consejería de Economía, Conocimiento, Empresas y Universidad (ref. P18-RT-4455); InFLAMES Flagship Programme of the Academy of Finland (decision number: 337530), Grant FJC2020-044453-I funded by MCIN/AEI/10.13039/501100011033 and by “European Union NextGenerationEU/PRTR”.

## Disclosure Statement

The authors declare no conflicts of interest relevant to this manuscript.

## Data and Code Availability

All analysis scripts and algorithm implementations are available in the open-source GGIR R package and at https://github.com/jhmigueles/spt_detection_hip.

## Notes

### Competing Interest Statement

The authors have declared no competing interest.

### Author Declarations

Ethics Committee of the University of Granada gave ethical approval for this work

