## Supplementary Material for "Estimating the sleep period time window based on a hip-worn accelerometer collected in children and adults"

**3-fold nested cross-validation of hyperparameters for HDCZA**

*Table S1: Root Mean Square Error (RMSE) results across nested cross-validation folds for each hyperparameter bound set tested in the HDCZA algorithm. The bound set with the lowest aggregated RMSE (mean × standard deviation) is highlighted in grey.*

| Fold | Bound Set | RMSE | HDCZA_threshold | spt_min_block_dur | spt_max_gap_dur | spt_max_gap_ratio |
|---|---|---|---|---|---|---|
| 1 | 1 | 1.094 | 0.402 | 24 | 39 | 0.091 |
| 2 | 1 | 0.966 | 0.249 | 35 | 54 | 0.314 |
| 3 | 1 | 0.909 | 0.275 | 29 | 44 | 0.278 |
| 1 | 2 | 1.035 | 0.250 | 60 | 56 | 0.644 |
| 2 | 2 | 0.950 | 0.605 | 60 | 57 | 0.077 |
| 3 | 2 | 0.915 | 0.335 | 54 | 59 | 0.347 |

RMSE: root mean square error.

**Confusion Matrices for the Quintile-based Agreement for the SPT duration**

*Table S2: Confusion matrix for quintile classification of SPT duration (HDCZA vs. reference). Cells show n, row %. Weighted Cohen's kappa (squared) = 0.556, p <0.001. N = 519.*

| Reference quintile | Q1 | Q2 | Q3 | Q4 | Q5 |
|---|---|---|---|---|---|
| Q1 | 53, 50.0% | 24, 22.6% | 17, 16.0% | 2, 1.9% | 10, 9.4% |
| Q2 | 10, 10.2% | 45, 45.9% | 23, 23.5% | 7, 7.1% | 13, 13.3% |
| Q3 | 10, 9.3% | 9, 8.3% | 52, 48.1% | 17, 15.7% | 20, 18.5% |
| Q4 | 8, 7.8% | 3, 2.9% | 18, 17.5% | 36, 35.0% | 38, 36.9% |
| Q5 | 9, 8.7% | 5, 4.8% | 6, 5.8% | 14, 13.5% | 70, 67.3% |

*Table S3: Confusion matrix for quintile classification of SPT duration (HorAngle vs. reference). Cells show n, row %. Weighted Cohen's kappa (squared) = 0.575, p <0.001. N = 519.*

| Reference quintile | Q1 | Q2 | Q3 | Q4 | Q5 |
|---|---|---|---|---|---|
| Q1 | 56, 56.6% | 31, 31.3% | 5, 5.1% | 3, 3.0% | 4, 4.0% |
| Q2 | 13, 12.6% | 42, 40.8% | 30, 29.1% | 7, 6.8% | 11, 10.7% |
| Q3 | 7, 6.5% | 10, 9.3% | 45, 41.7% | 31, 28.7% | 15, 13.9% |
| Q4 | 9, 8.6% | 7, 6.7% | 10, 9.5% | 40, 38.1% | 39, 37.1% |
| Q5 | 14, 13.5% | 4, 3.8% | 7, 6.7% | 17, 16.3% | 62, 59.6% |

**Linear Mixed Models for the Effect of Confounders on Algorithms' performance**

*Table S4: Regression summary for the effect of confounders (i.e., age, sex, day type, and non-wear) on the estimation of sleep onset and wake-up time with the HDCZA and the HorAngle algorithms.*

| | | HDCZA | | | | HorAngle | | | |
|---|---|---|---|---|---|---|---|---|---|
| Outcome | Term | Estimate | Std. Error | t Value | P Value | Estimate | Std. Error | t Value | P Value |
| Sleep Onset | Intercept | -0.133 | 0.197 | -0.674 | 0.501 | 0.168 | 0.173 | 0.972 | 0.333 |
| | Age group (children) | 0.214 | 0.217 | 0.986 | 0.326 | 0.332 | 0.187 | 1.772 | 0.079 |
| | Sex (female) | 0.216 | 0.202 | 1.067 | 0.289 | 0.156 | 0.174 | 0.896 | 0.373 |
| | Weekend day | 0.312 | 0.162 | 1.919 | 0.056 | -0.026 | 0.156 | -0.165 | 0.869 |
| | Wrist non-wear (low) | -0.262 | 0.260 | -1.008 | 0.314 | -0.412 | 0.247 | -1.665 | 0.096 |
| | Wrist non-wear (moderate) | 0.110 | 0.398 | 0.277 | 0.782 | -0.323 | 0.377 | -0.856 | 0.392 |
| | Wrist non-wear (high) | 0.147 | 0.549 | 0.268 | 0.789 | -0.875 | 0.517 | -1.690 | 0.092 |
| | Hip non-wear (low) | -0.136 | 0.258 | -0.526 | 0.599 | -0.015 | 0.246 | -0.061 | 0.951 |
| | Hip non-wear (moderate) | -0.320 | 0.385 | -0.831 | 0.406 | -0.207 | 0.367 | -0.564 | 0.573 |
| | Hip non-wear (high) | -0.447 | 0.520 | -0.859 | 0.391 | -0.049 | 0.491 | -0.099 | 0.921 |
| | Non-wear mismatch | -0.139 | 0.254 | -0.547 | 0.584 | **-0.740** | **0.240** | **-3.088** | **0.002 **** |
| Wake-up | Intercept | **0.372** | **0.142** | **2.613** | **0.009 **** | 0.167 | 0.180 | 0.926 | 0.356 |
| | Age group (children) | **-0.366** | **0.151** | **-2.420** | **0.016 *** | -0.212 | 0.194 | -1.093 | 0.277 |
| | Sex (female) | **-0.419** | **0.139** | **-3.024** | **0.003 **** | -0.187 | 0.179 | -1.048 | 0.297 |
| | Weekend day | 0.163 | 0.145 | 1.128 | 0.260 | -0.035 | 0.172 | -0.205 | 0.838 |
| | Wrist non-wear (low) | 0.377 | 0.224 | 1.685 | 0.093 | 0.071 | 0.269 | 0.266 | 0.791 |
| | Wrist non-wear (moderate) | 0.556 | 0.340 | 1.633 | 0.103 | 0.167 | 0.409 | 0.409 | 0.683 |
| | Wrist non-wear (high) | 0.525 | 0.464 | 1.132 | 0.258 | **-1.337** | **0.560** | **-2.387** | **0.017 *** |
| | Hip non-wear (low) | -0.355 | 0.223 | -1.590 | 0.112 | 0.067 | 0.268 | 0.250 | 0.802 |
| | Hip non-wear (moderate) | **-0.848** | **0.334** | **-2.542** | **0.011 *** | -0.469 | 0.399 | -1.174 | 0.241 |
| | Hip non-wear (high) | **-1.698** | **0.441** | **-3.847** | **<0.001 ***** | -0.057 | 0.532 | -0.107 | 0.915 |
| | Non-wear mismatch | 0.348 | 0.214 | 1.622 | 0.105 | -0.441 | 0.259 | -1.703 | 0.089 |

### Visualisation of example nights

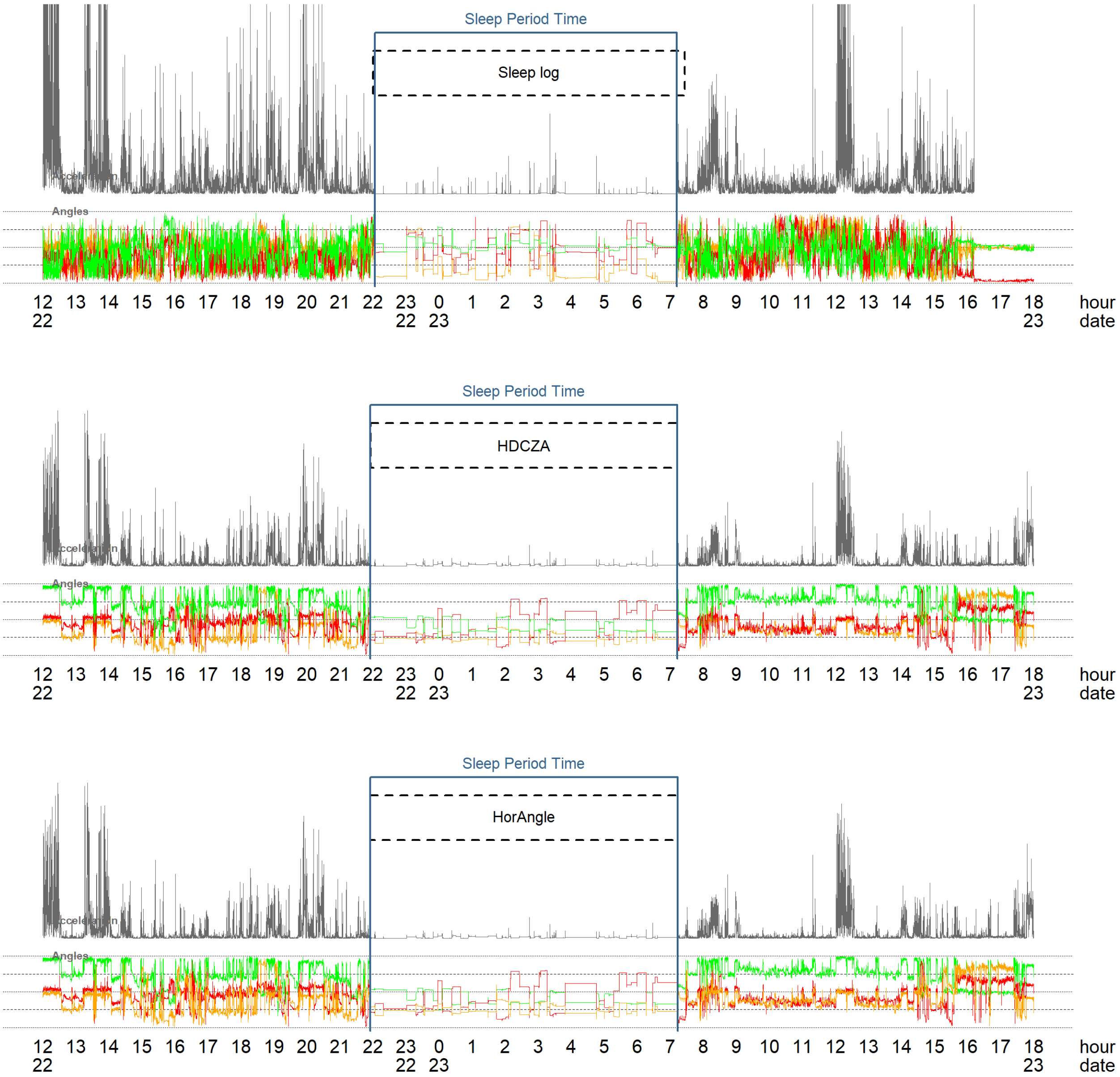


*Figure S1: Classification of a example night guided by sleeplog in wrist data, and by HDCZA and HorAngle in hip data. In this example, both algorithms show a similar performance compared to the reference method.*

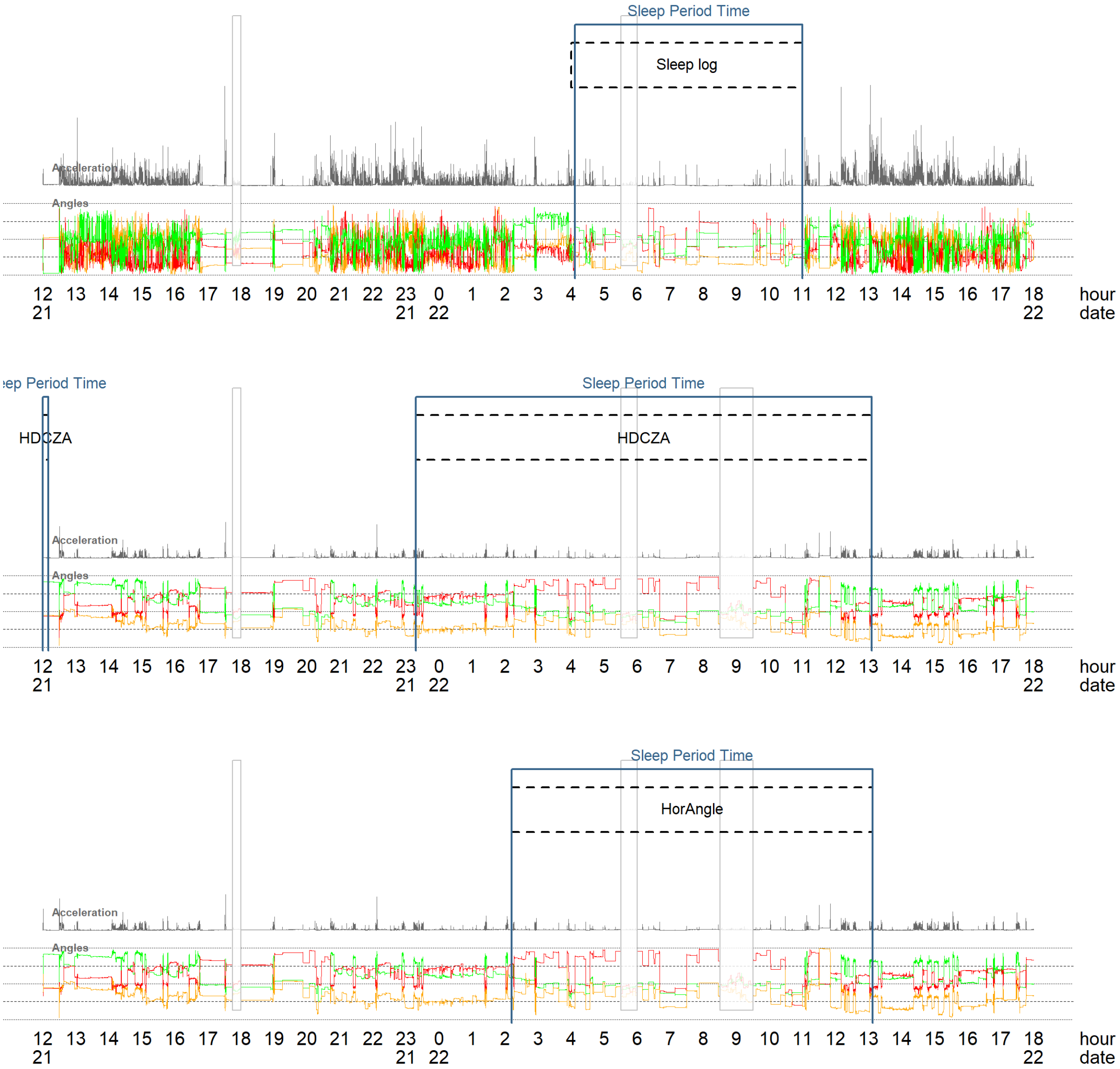


*Figure S2: Classification of an example night guided by sleeplog in wrist data, and by HDCZA and HorAngle in hip data. In this example the HorAngle estimates are closer to the reference method than the HDCZA algorithm.*

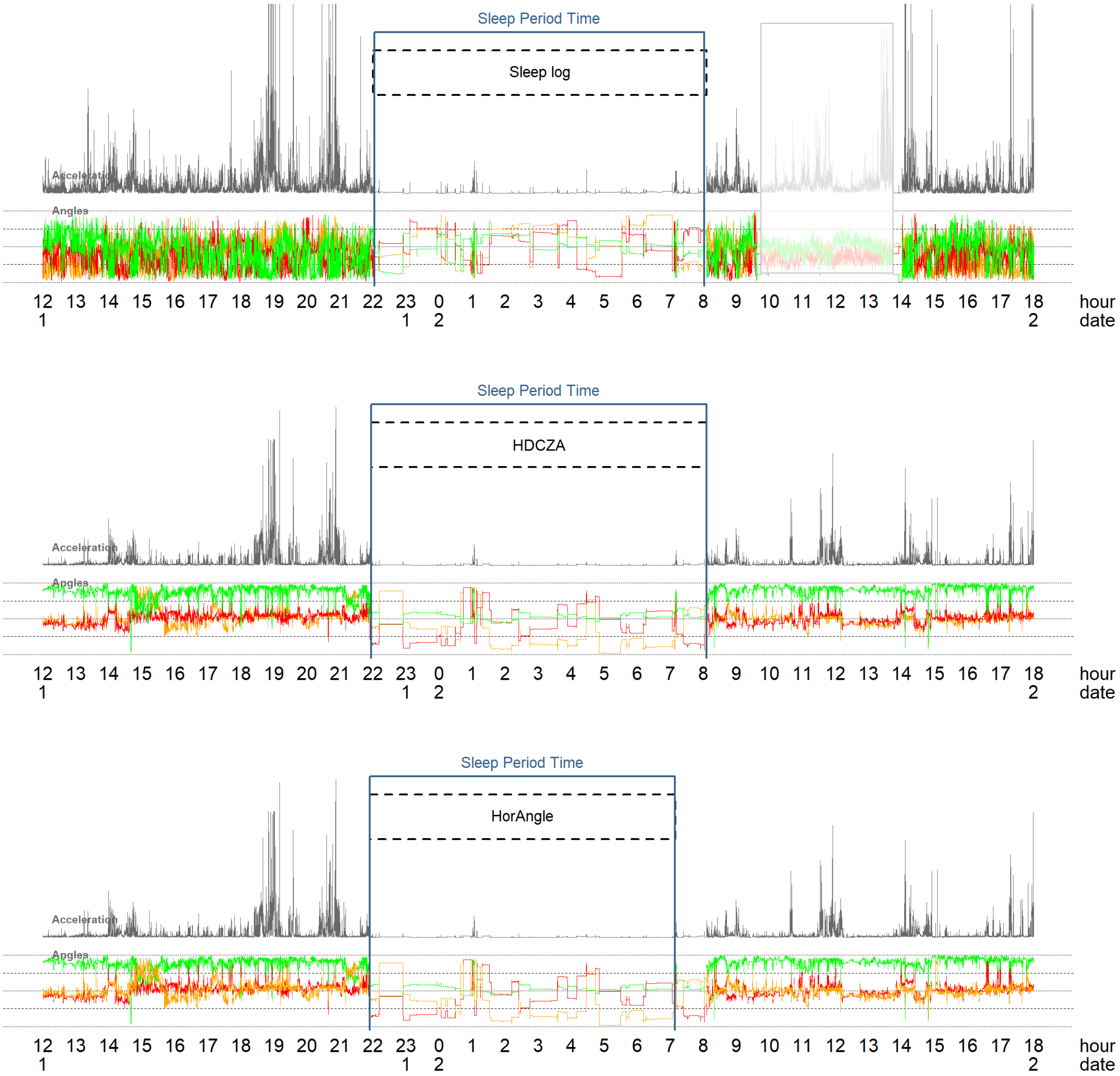


*Figure S3: Classification of a example night guided by sleeplog in wrist data, and by HDCZA and HorAngle in hip data. In this example the HorAngle estimates wakeup time earlier than the reference method, while HDCZA seems to agree better with the reference method.*

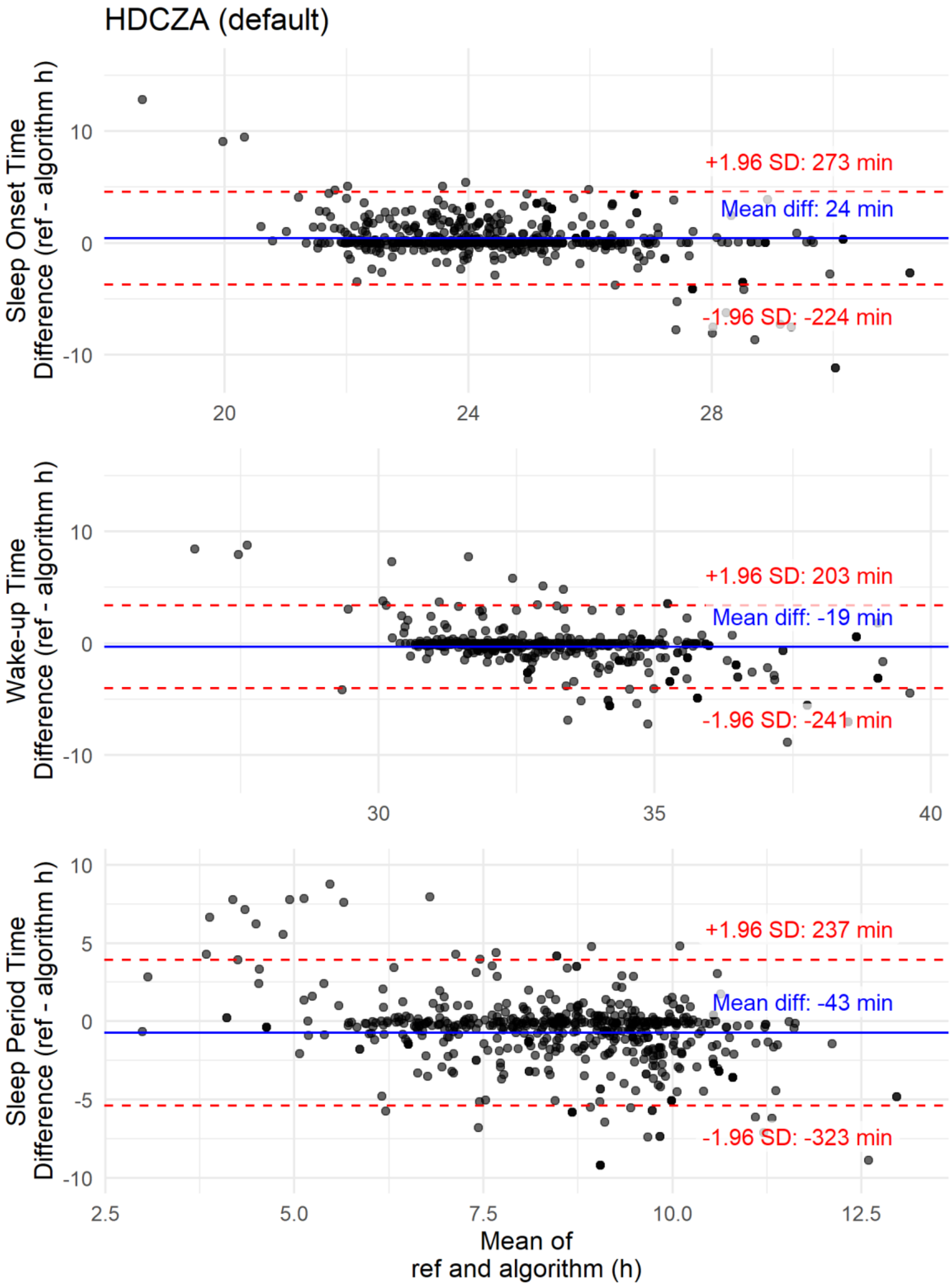


*Figure S4: Bland Altman plots for the comparison of default HDCZA estimated sleep onset time, wake-up time, and SPT with reference values.*

**GGIR example calls applying the optimised parameters**

```
# HDCZA optimised parameters

GGIR(
  # SPT algorithm specification
  HASPT.algo = "HDCZA",

  # settings to ensure that guider window is the final SPT window
  HASIB.algo = "vanHees2015",
  anglethreshold = 100,
  timethreshold = 1,
  sib_must_fully_overlap_with_TimeInBed = c(FALSE, FALSE),
  relyonguider = TRUE,

  # Optimised parameters
  HDCZA_threshold = 0.259,
  spt_min_block_dur = 34,
  spt_max_gap_dur = 52,
  spt_max_gap_ratio = 0.282,
)


# HorAngle optimised parameters

GGIR(
  # SPT algorithm specification
  HASPT.algo = "HorAngle",
  sensor.location = "hip",

  # settings to ensure that guider window is the final SPT window
  HASIB.algo = "vanHees2015",
  anglethreshold = 100,
  timethreshold = 1,
  sib_must_fully_overlap_with_TimeInBed = c(FALSE, FALSE),
  relyonguider = TRUE,

  # Optimised parameters
  HorAngle_threshold = 58,
  spt_min_block_dur = 60,
  spt_max_gap_dur = 20,
  spt_max_gap_ratio = 0.034,
)
```
